# Characteristics of COVID-19 Clinical Trials in India Based on the Registration Data on CTRI (Clinical Trials Registry- India): a cross-sectional analysis

**DOI:** 10.1101/2020.10.14.20212761

**Authors:** P Sathiyarajeswaran, MS Shree Devi, K Kanakavalli, NP Vinod

**Affiliations:** Siddha Central Research Institute (CCRS), Chennai, Tamilnadu, India; Central Council for Research in Siddha, Ministry of AYUSH, Chennai, India

**Keywords:** COVID-19, CTRI, Clinical Trial Registry of India, Ministry of AYUSH, Clinical trial, Interventional, Randomized, Blinding

## Abstract

**Objectives:** The 2019 pandemic of coronavirus disease (COVID-19) has prompted several efforts to find safe and effective drugs, but little is understood as to where early efforts were centered. Several clinical trials, both Allopathy and AYUSH medicines have been registered in the Clinical Trial Registry of India (CTRI). We aimed to characterize and extract relevant data registered under CTRI for COVID-19.

**Materials and Methods:** A cross-sectional analysis was performed of clinical trials for the treatment of COVID-19 that were registered in the Clinical Trial Registry of India (CTRI) from 1^st^ March 2020 to 22^nd^ June 2020. Relevant trial records were downloaded, deduplicated, and independently analyzed by three reviewers.

**Main outcomes:** Trial intervention, design, sponsorship, phase of the trial, and indicated outcomes.

**Results:** 233 COVID-19 clinical trials, was registered from India in CTRI. Out of these, 146 were Interventional trials, 84 Observational trials, and three Post-marketing surveillance. Questionnaire and survey-based intervention occupy a significant portion. Randomized control trials are large in number 37.8% than non-randomized. 20% of the trials were recruiting patients, and the Research institution (34%) sponsored more than half of the trials. Global trials are minimal, occupying 3% of total trials and Indian trials were 97%. In most of the trials, the interventional agent is either multiple drug combinations or compound drug formulations compared to single drug administration. Among the trials, 46 Allopathic interventions, 41 Ayurveda interventions, 14 Homeopathy interventions, one in Unani, and 2 in yoga and Naturopathy.

**Conclusion:** This study will provide a background of COVID-19 clinical trials registered in CTRI and provide specific issues observed related to clinical trial designs, which offer information to perform clinical trials on COVID-19

## Introduction

Since Coronavirus Disease 2019 (COVID-19) [1] emerged from Wuhan, China, in December 2019, has become pandemic and rapidly crossed all countries by reporting cases by April 2020[2]. There are 4 10 461 cases and 13 254 deaths in India as of 30th June 2020. These data correspond to the imminent risk facing the country. Transmission of the virus spreads via droplets, physical contact with infected individuals, contaminated surfaces [3]. Fever, Cough, Headache, and Throat pain are the most common symptoms [4]. The severe infection leads to pneumonia, acute respiratory distress syndrome (ARDS), and sometimes multi-organ failures such as kidney failure and even death [5]. Coronaviruses (CoVs) are the family of Coronaviridae with four gene era (alpha, beta, gamma, and delta), and only the alpha and beta-strains identified to be pathogenic to human [6, 7]. There are no specific anti-viral drugs to treat COVID-19. As of now, symptomatic supportive therapy will be the treatment protocol. Govt of India has released guidelines via the Ministry of Health and via the AYUSH Ministry for mitigation of COVID-19. However, medical researchers on both the ends are rigorously working for solutions to combat COVID-19. Ethical issues involved in these attempts should guide by existing Meuri guidelines that evolved during the Ebola outbreak.

In this context, it becomes mandatory to analyze the conduct, creditability, ethical issues, willingness, consensual integration; intervention used out of pocket expenses, availability of the drugs, and drugs that emerged out of Indian origin. Above all, the characters were extracted from the trials registered in Clinical Trials Registry—India (CTRI).

The Clinical Trials Registry—India (CTRI) is a free and online public record domain for registration of clinical trials conducted in India from 20th July 2007. Drug Controller General of India has made it mandatory to register clinical trials at CTRI since 15th June 2009. Registering clinical trials is a contemporary issue in present health researchers [8]. The analysis performed using the data available online with CTRI helped us look for the characters mentioned above in the purview of COVID-19, which may help more in future pandemics. In the present study, the content from clinical trials registered in India was summarized and analyzed from various perspectives and design of clinical trials for COVID-19.

## Materials and Methods

### Search Strategy

‘COVID-19’ and ‘INDIA’ ‘Coronavirus’ or ‘COVID-19’ or ‘COVID19’ or ‘2019 novel coronavirus’ or ‘2019-nCoV’ or ‘SARS-CoV-2’ (were used as the keywords to search for all COVID-19 clinical trials registered from India on CTRI (Clinical Trials Registry – India) [9] from 1^st^ March 2020 to 22^nd^ June 2020. All retrieved records were downloaded. The following data were collected: study title, study type, study design, primary outcome measures, condition, Number of groups, Intervention, blinding, phase of trials, and sample size. A standard Microsoft-Excel database was created for the analysis.

### Trial selection

We included all studies conducted on patients diagnosed with COVID-19. First, we selected clinical trials based on the ‘study type’ variable. The variable contains interventional and observational studies and others. We included all AYUSH system trials, and our study does not have any limits based on the outcome.

### Statistical Analyses

All continuous values were expressed as a median and interquartile range, and categorical variables are expressed as percentages. All statistical procedures were performed using the SPSS software (Version 26).

### Data extraction and management

We extracted the following information from each trial: CTRI trial number, registration date, recruitment status (recruiting, not yet recruiting, withdrawn or canceled), recruitment country, phase (0, 1, 2, 3, 4, 1–2, 2–3, not applicable and missing), sponsor, health condition, an intervention model (single-arm, parallel, cross-over, factorial, platform trial or sequential), type of study, study design, sample size, trial duration, allocation, blinding (open, single, double, triple or quadruple), primary outcomes.

## Results

The Number of COVID-19 Clinical Trials Registered in India, 233 COVID-19 clinical trials, was registered from India in CTRI. After the general analysis, 146 Interventional trials, 84 Observational trials, and 3 Post-marketing surveillance were registered. The rates of trial registration are incoherence, the incidence, and the intensity of the disease. There is an ascending pattern in registration from March reaching its peak in May and slowing down in June. India had registered 107 more trials than China in a set period of three months (n-India (233) - n-China (126) = n=107) (Table – 1)(Figure – 1,2)

**Figure: 1.**
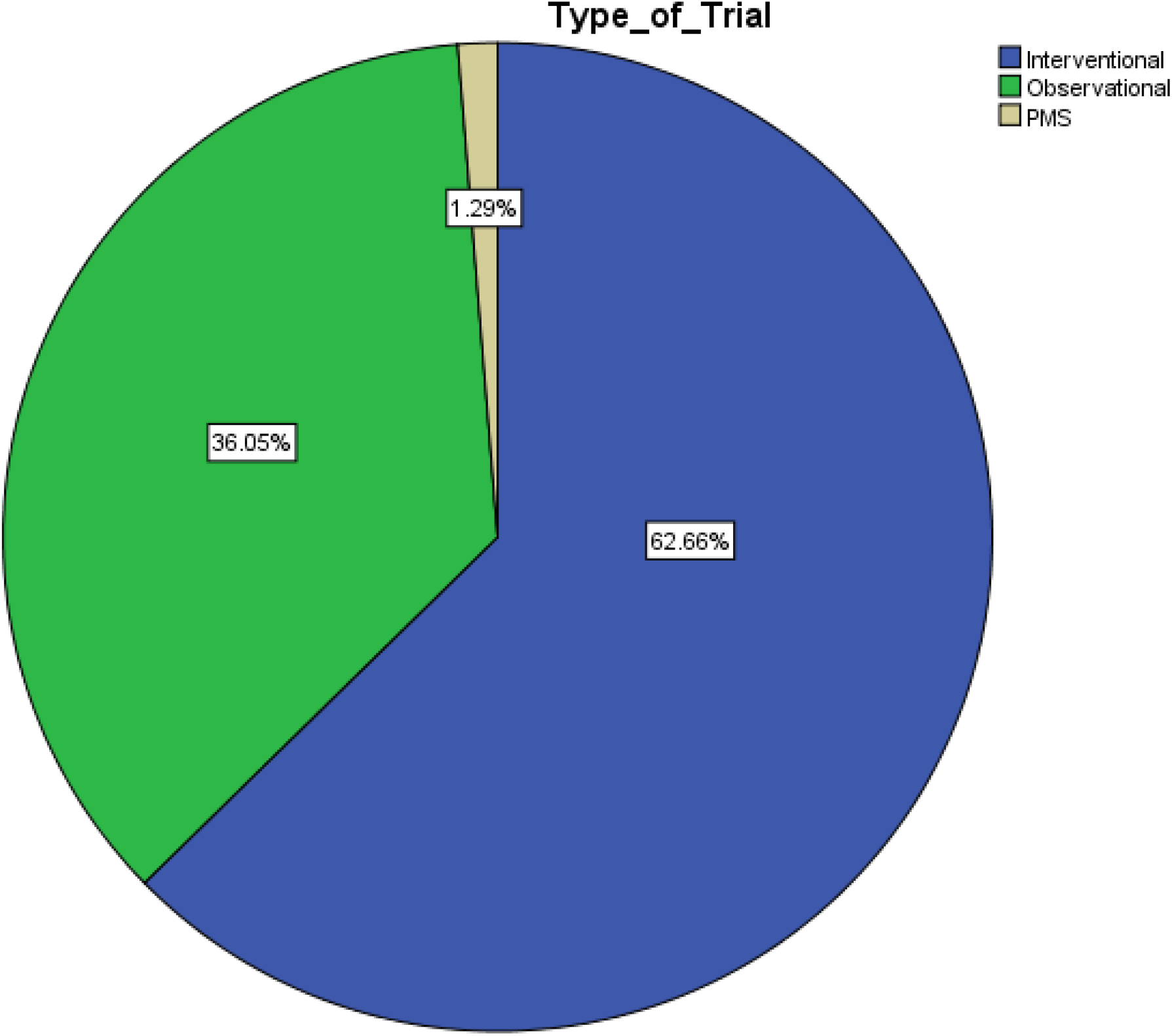
Type of Trial.

**Figure: 2.**
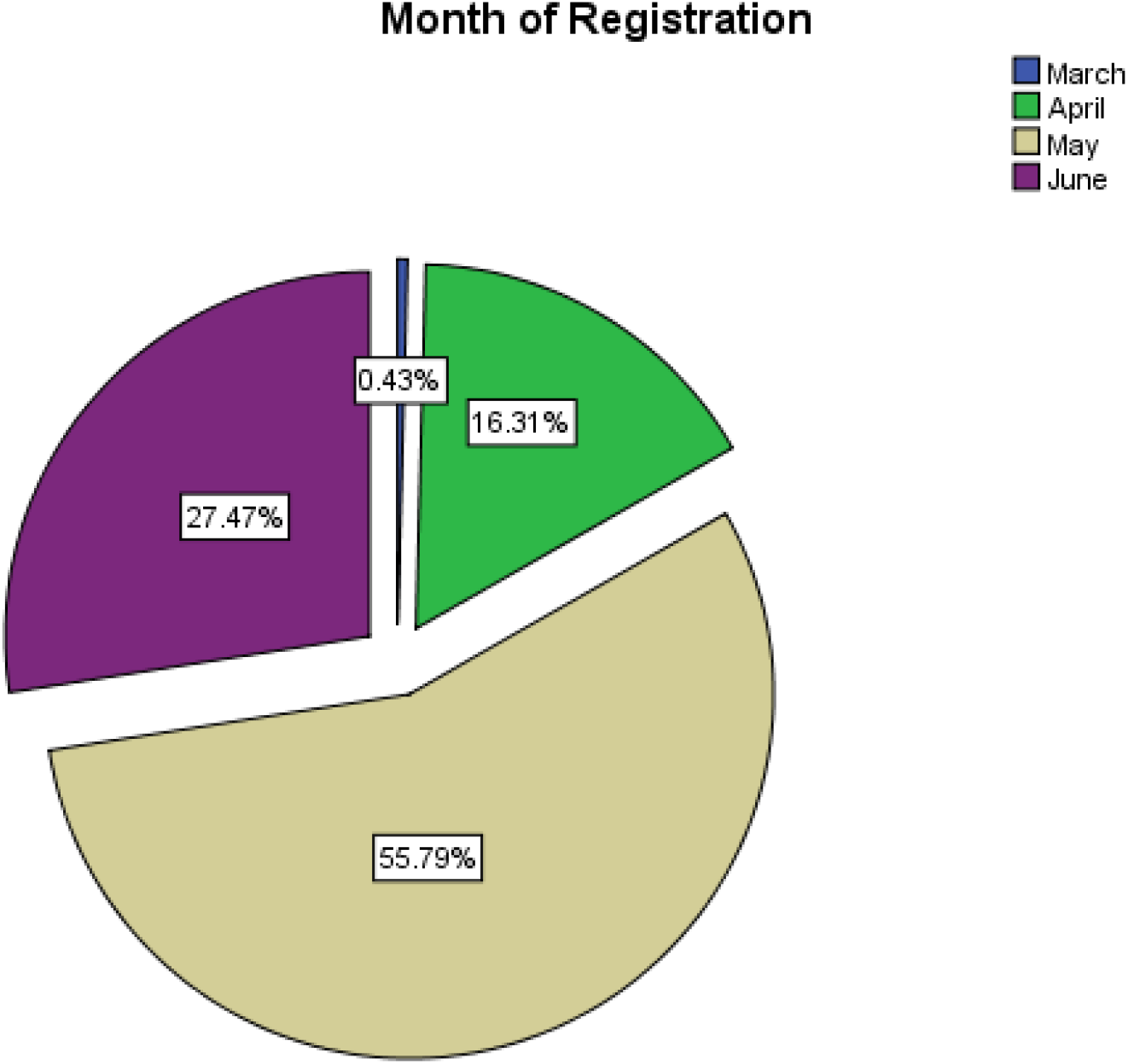
Month of Registration.

**Table 1.**
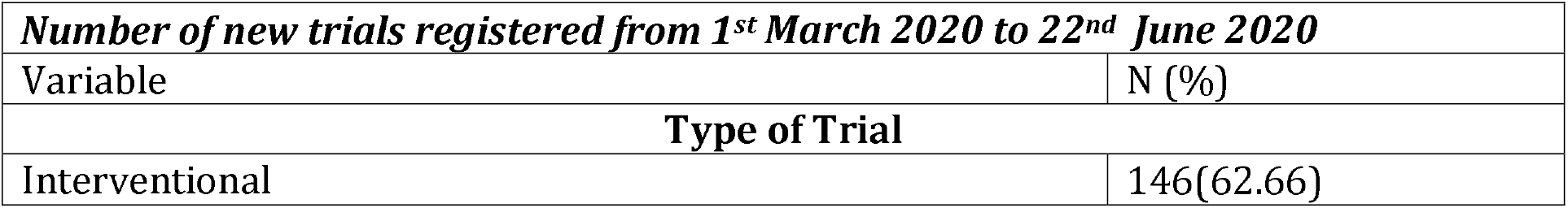
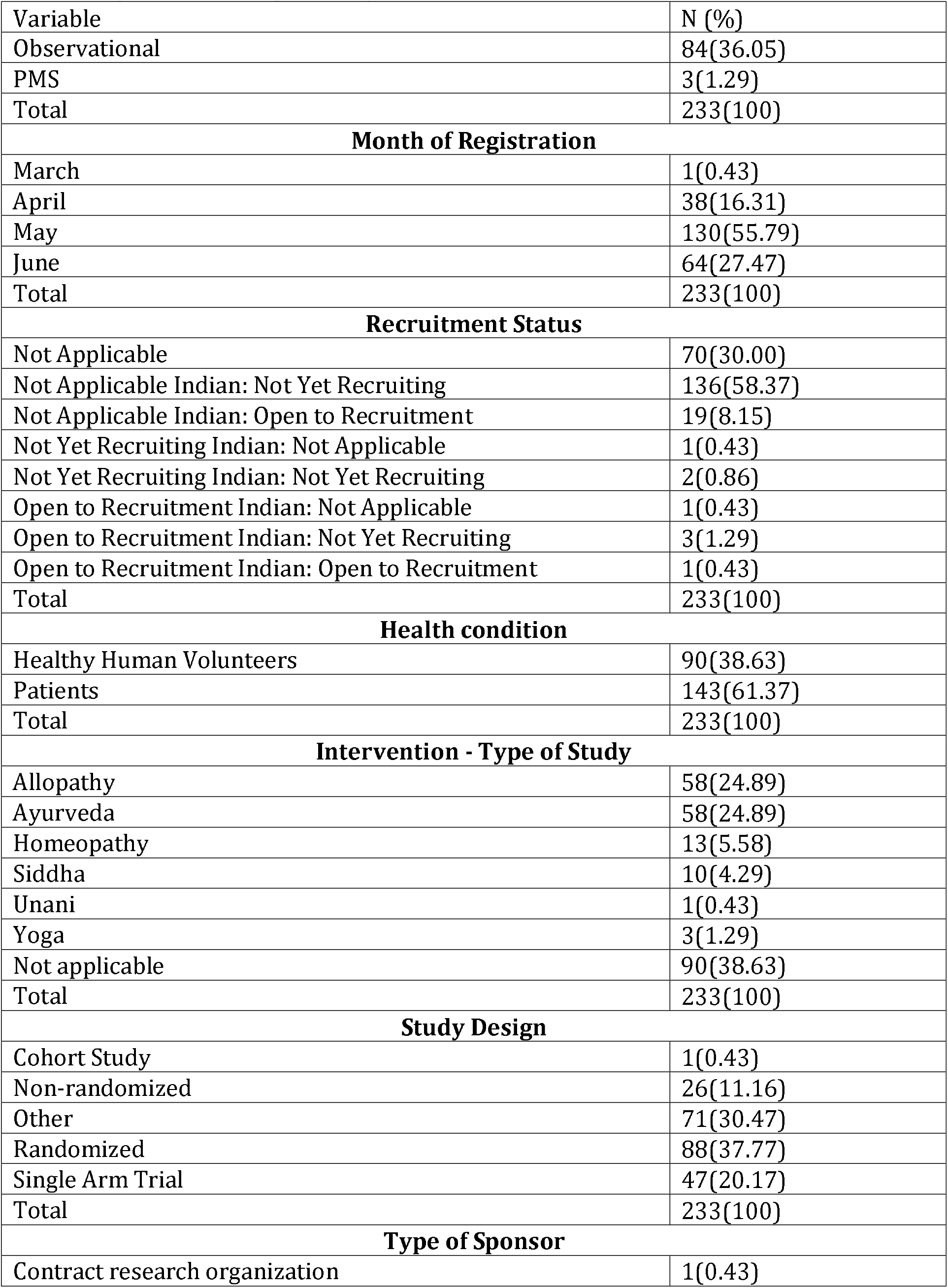
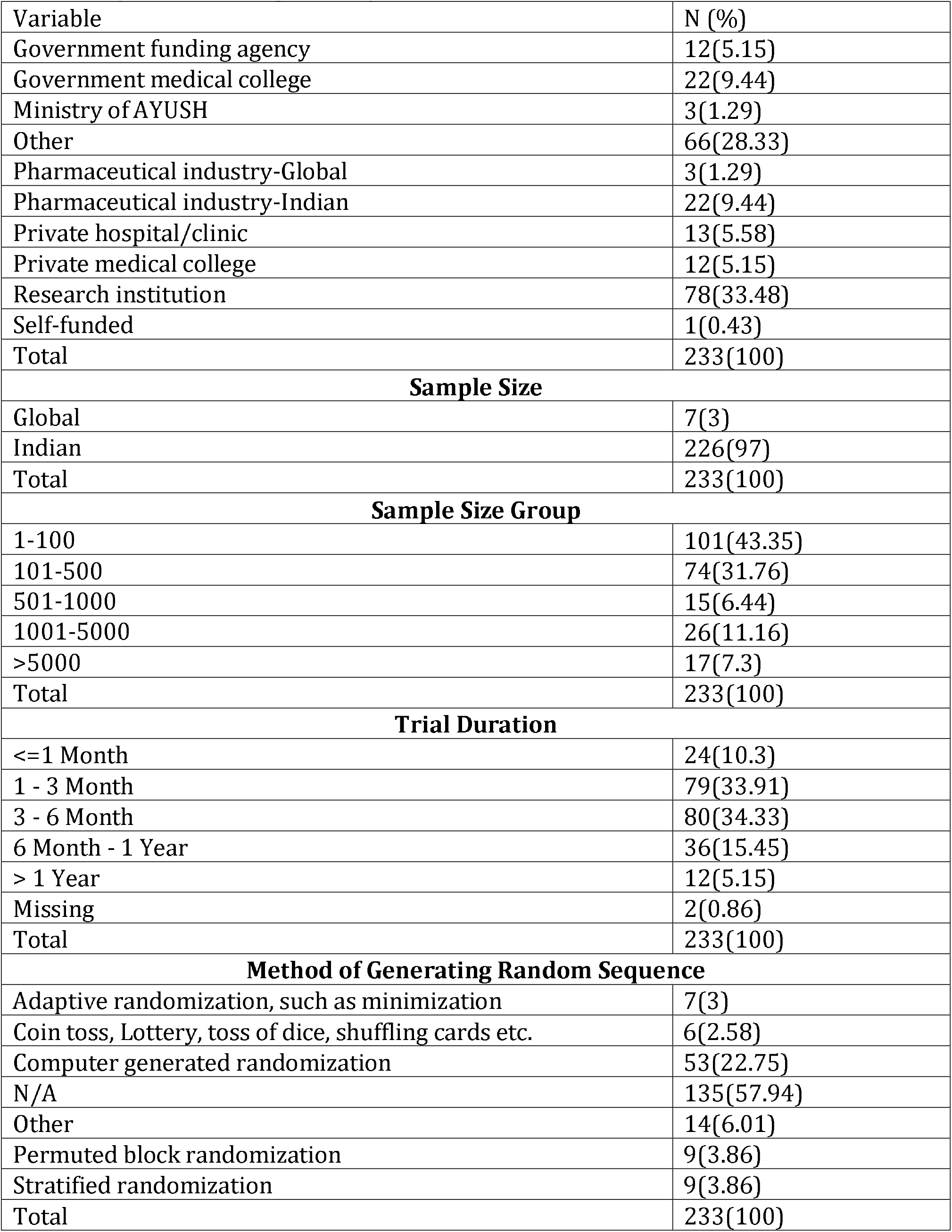
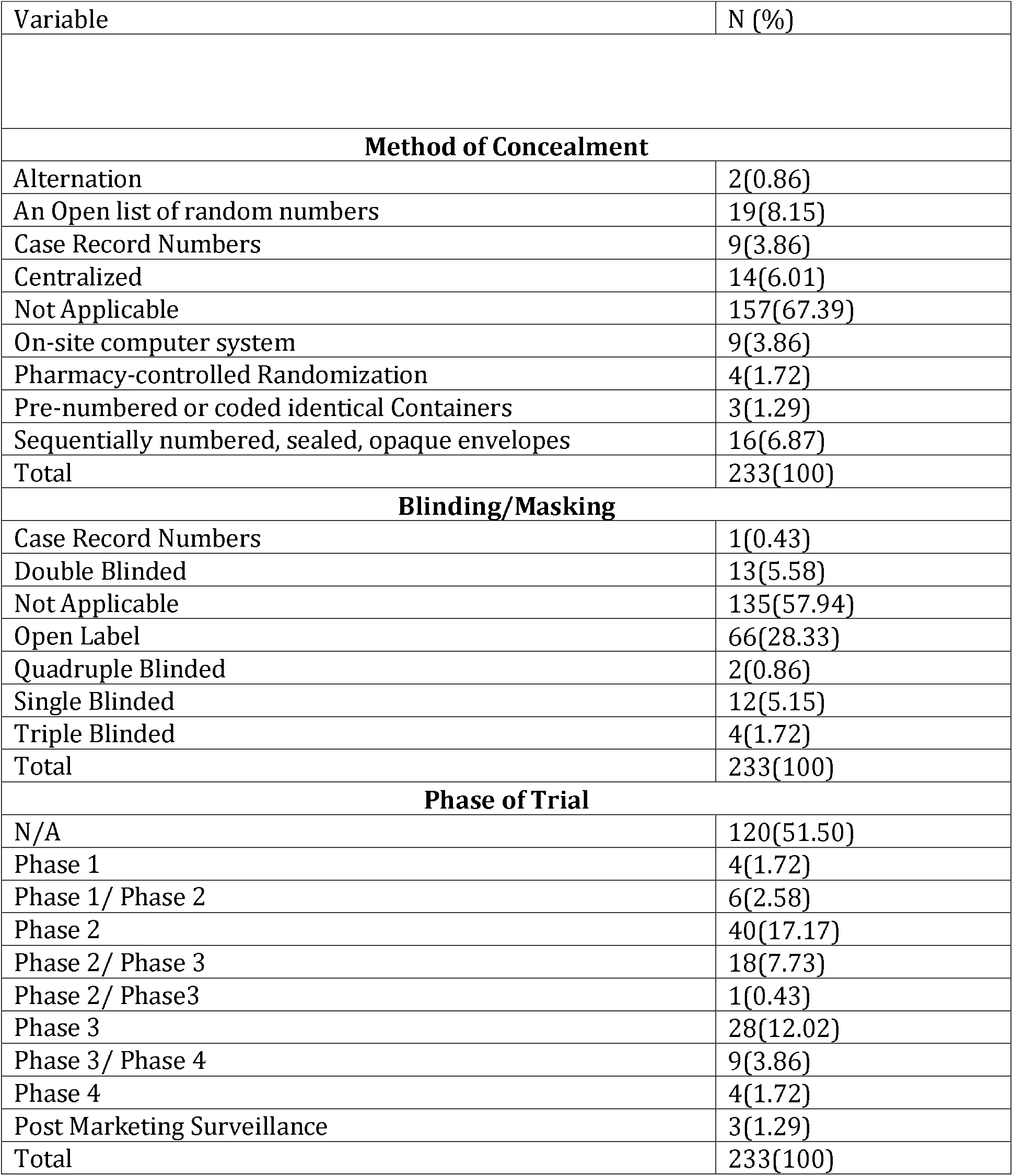
General characteristic of registered trials.

The recruitment status has been calculated here at least fifteen days after the trial is registered in CTRI. At this point, not less than 50 % of the trials have not started recruiting. COVID-19 is a peculiar condition wherein prophylaxis plays an important role, which is evident from 32% of the people either in the survey or as a prophylactic intervention have been registered under the category as healthy volunteers. However, it is also noteworthy that 62% of the trials are COVID-19 affected individuals, which may benefit human society by and large (Figure -3)

**Figure: 3.**
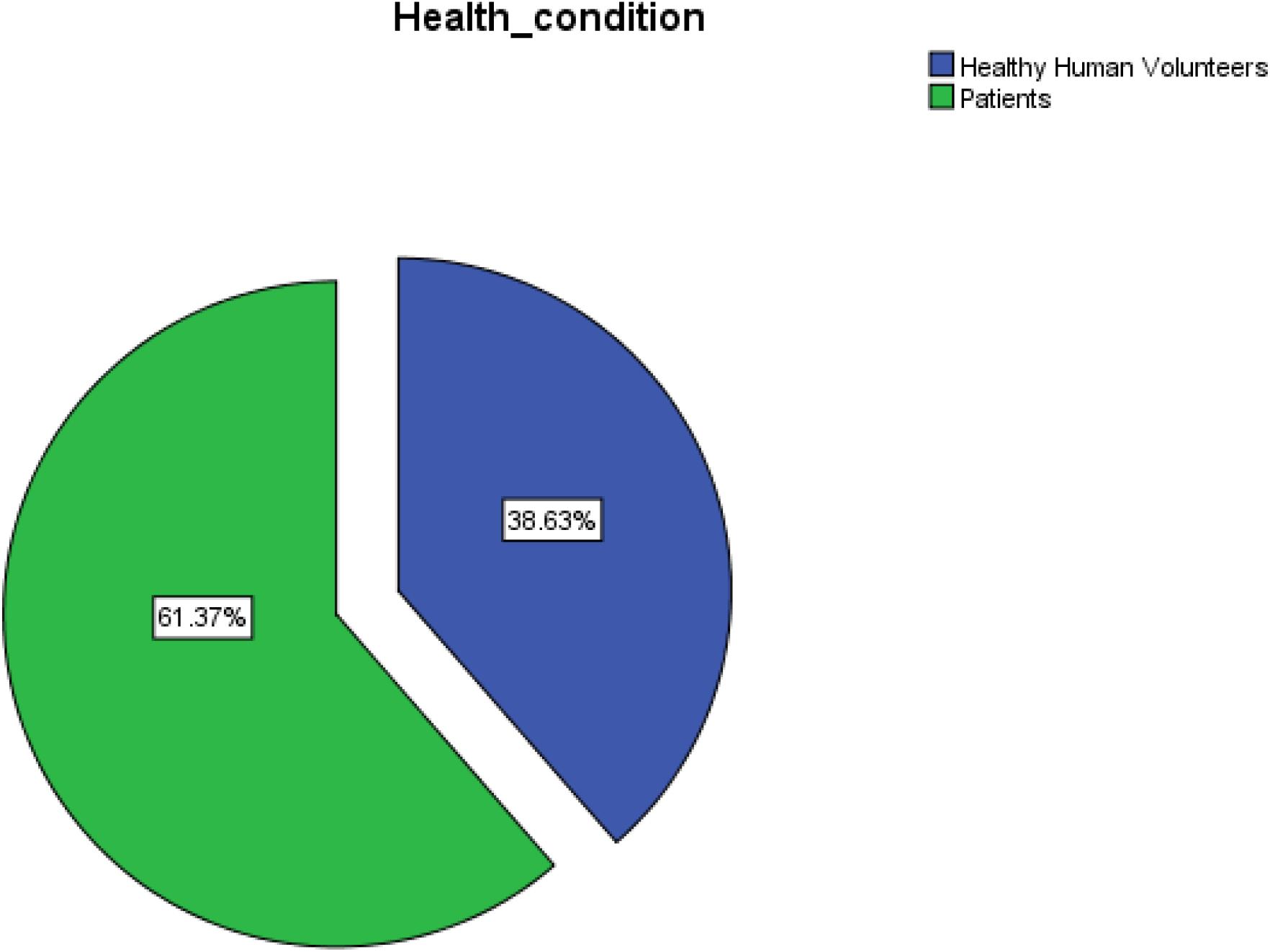
Health Condition.

The intervention of various Medical systems are described in descending order; surveys are dealt with separately. Allopathy and Ayurveda have a significant share of (25% each) followed by Homeopathy 5.6%, Siddha 4.3%, Unani 0.5%, Yoga 1.3% (Table – 1) (Figure -4). Questionnaire and survey-based intervention occupy a significant portion. There is a demand for analyzing the nature of people’s disease, psychological, social, and lifestyle adaptation toward the new regular life pattern in lockdown. In certain hospitals, integrated treatment is available for Ayurveda and Homeopathy, Ayurveda, and Yoga & Naturopathy. There are database observations reported in certain hospitals that portrayed integrated treatment between Allopathy and Siddha in Tamil Nadu. Clinical trials with integrated approaches of Allopathy and Siddha are happening at Noida. New Delhi.

**Figure: 4.**
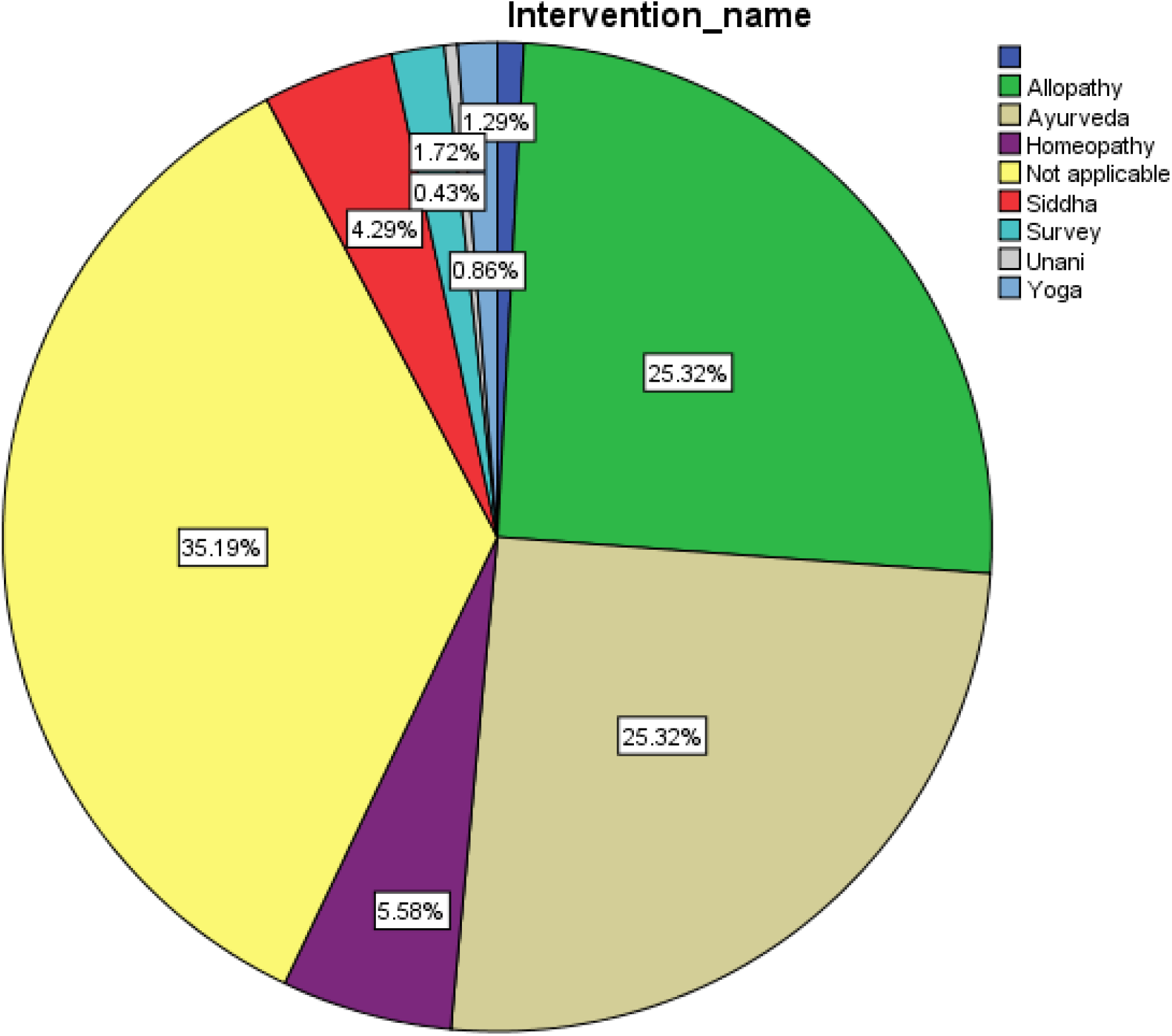
Intervention - Type of Study.

As of the current date, more than 46 Allopathic interventions from Anti-viral, Antimalarial, oncologic drugs, Immunotherapies, Vaccines, and Plasma therapy are in testing. At the same time, Siddha has 10 Medicines in the clinical trial platform. Ayurveda has used 41 medicines, 14 are used in Homeopathy Medicine, and Unani had one, and 2 in Yoga and Naturopathy were tried (Table -2).

**Table 2.**
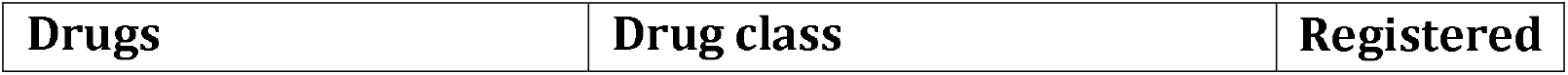
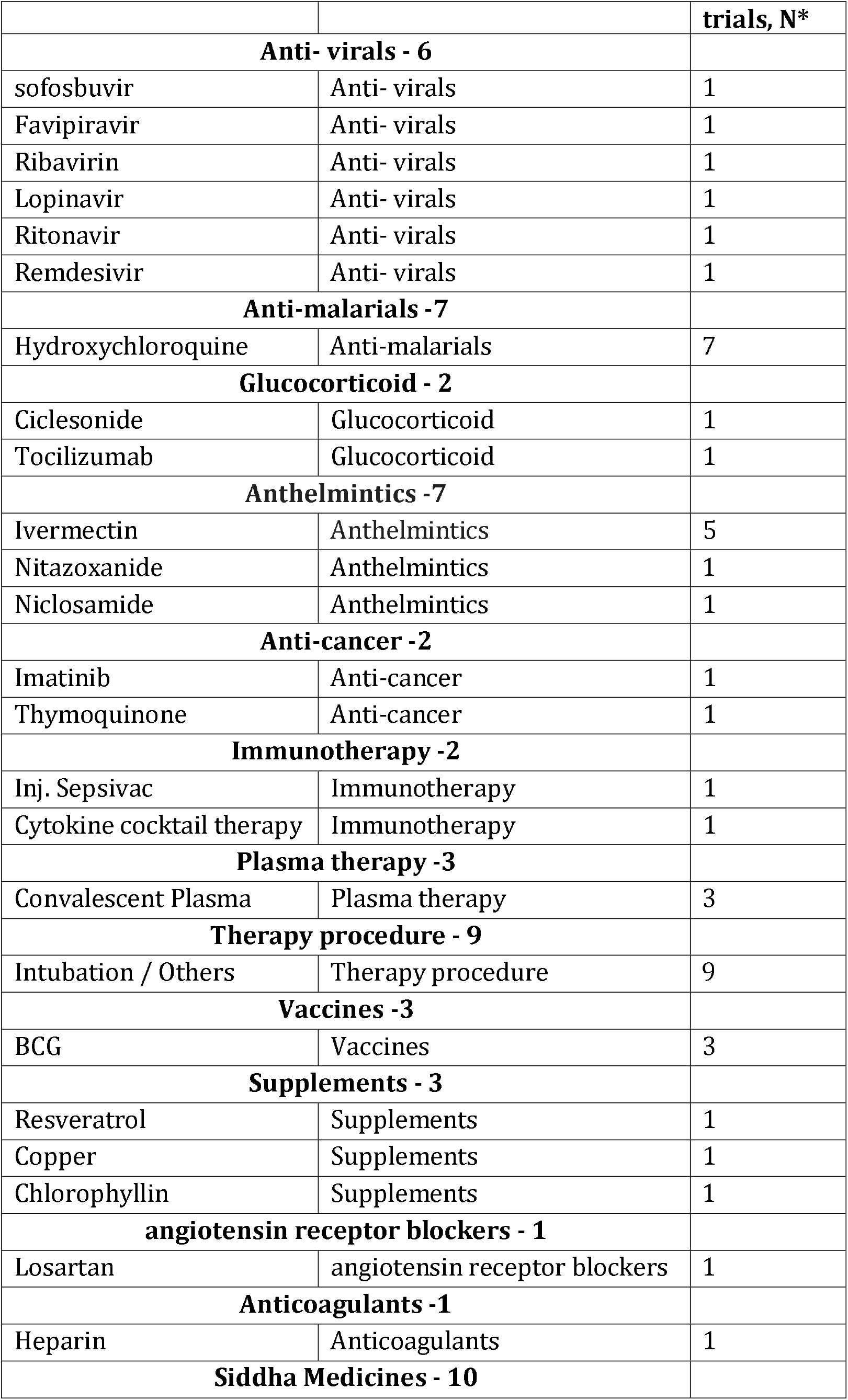
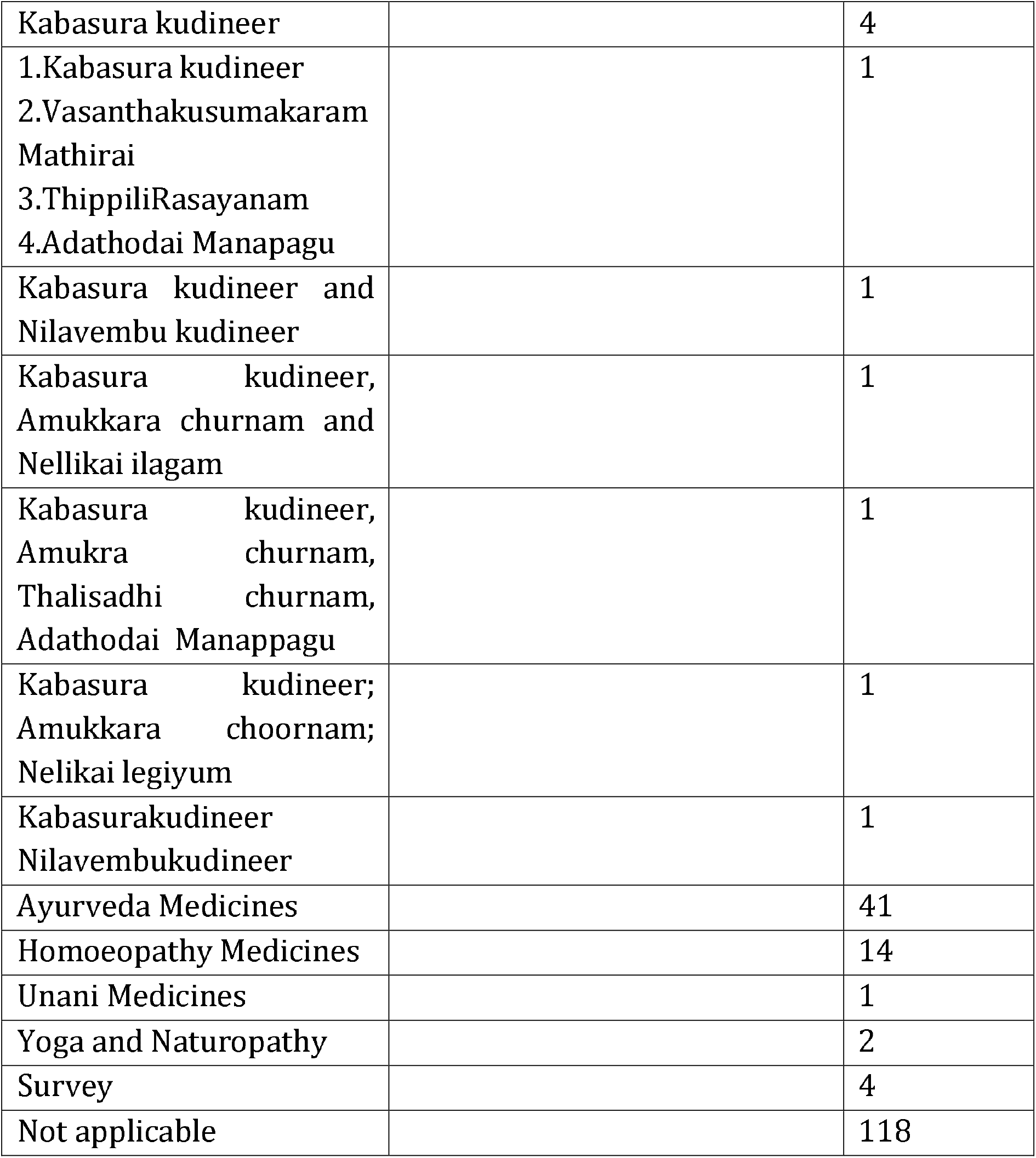
Products being assessed in Clinical Trial Registry of India (CTRI) for COVID-19 infection.

### Study design

Randomized control trials are large in number 37.8% than non-randomized, 11.2%, and Single-arm trials are next to RCT sharing 20.2%. Cohort studies are lesser 0.5% alone. Among the global trials except one study checking the efficacy of remdesiver and all other studies are observations and surveys. However, in the domestic trial scenario is a little different. Nearly 20% of the trials were recruiting patients, and more than half of the trials were sponsored by Research institution (34%) followed by others (29%), colleges (10%), Pharmaceutical industry – Indian (10%), and private colleges/ clinic (10%) (Table – 1) (Figure -5). The Number of COVID-19 clinical trials registered in CTRI is considerably high over the past three months.

**Figure: 5.**
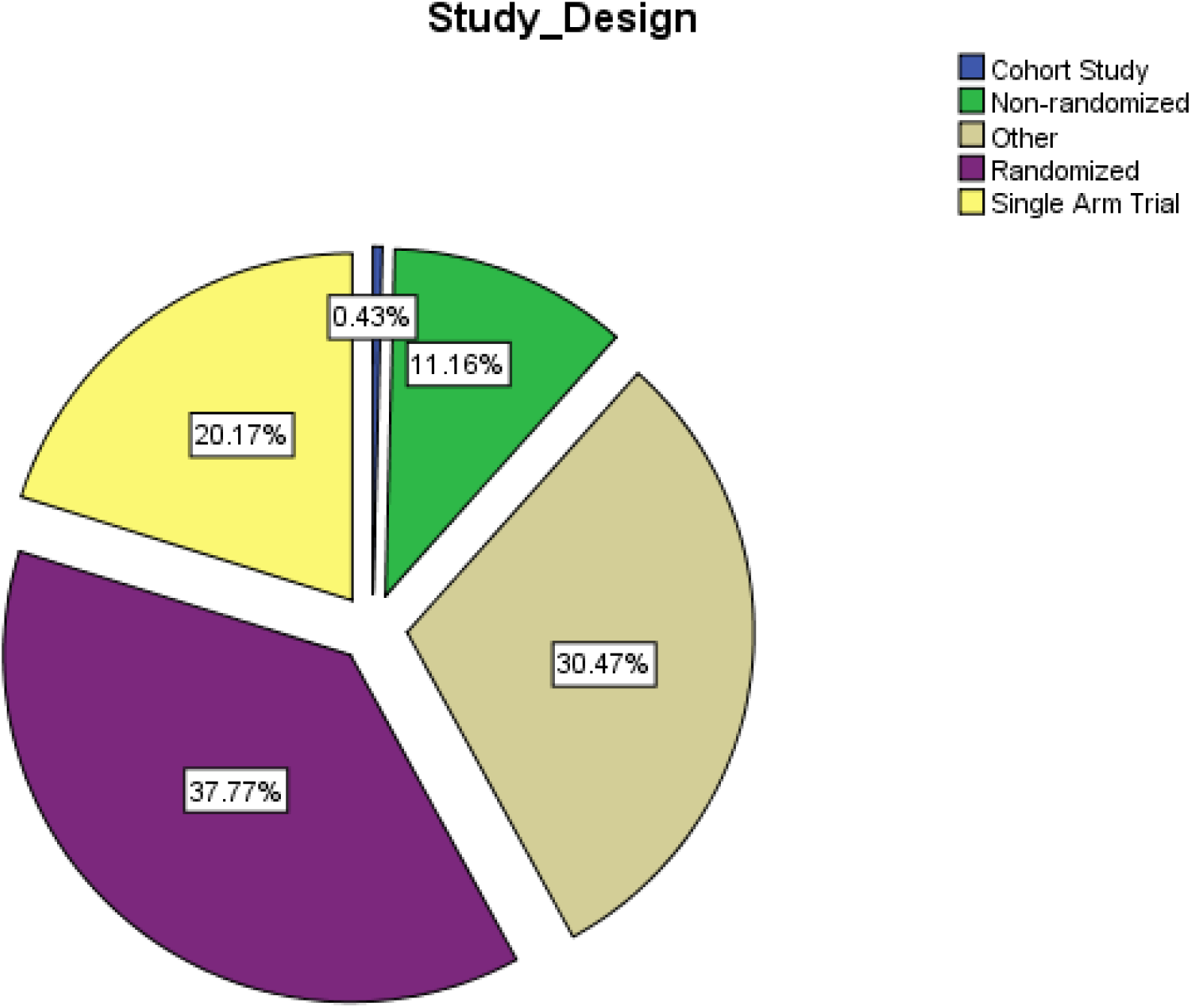
Study Design.

### Sponsors

As in other scenarios, Research Institution occupies Trials’ significant share (33%), followed by Pharmaceutical Industries and Medical colleges. Private hospitals and medical colleges register 25 %of trials (Table – 1).

### Percentage of Global trials

Intercontinental of the inter-country collaborated global trial is minimal in Number, indicating the complexity of the disease. Global trials are minimal, occupying 3% of total trials and Indian trials were 97% (Table – 1) (Figure -6).

**Figure: 6.**
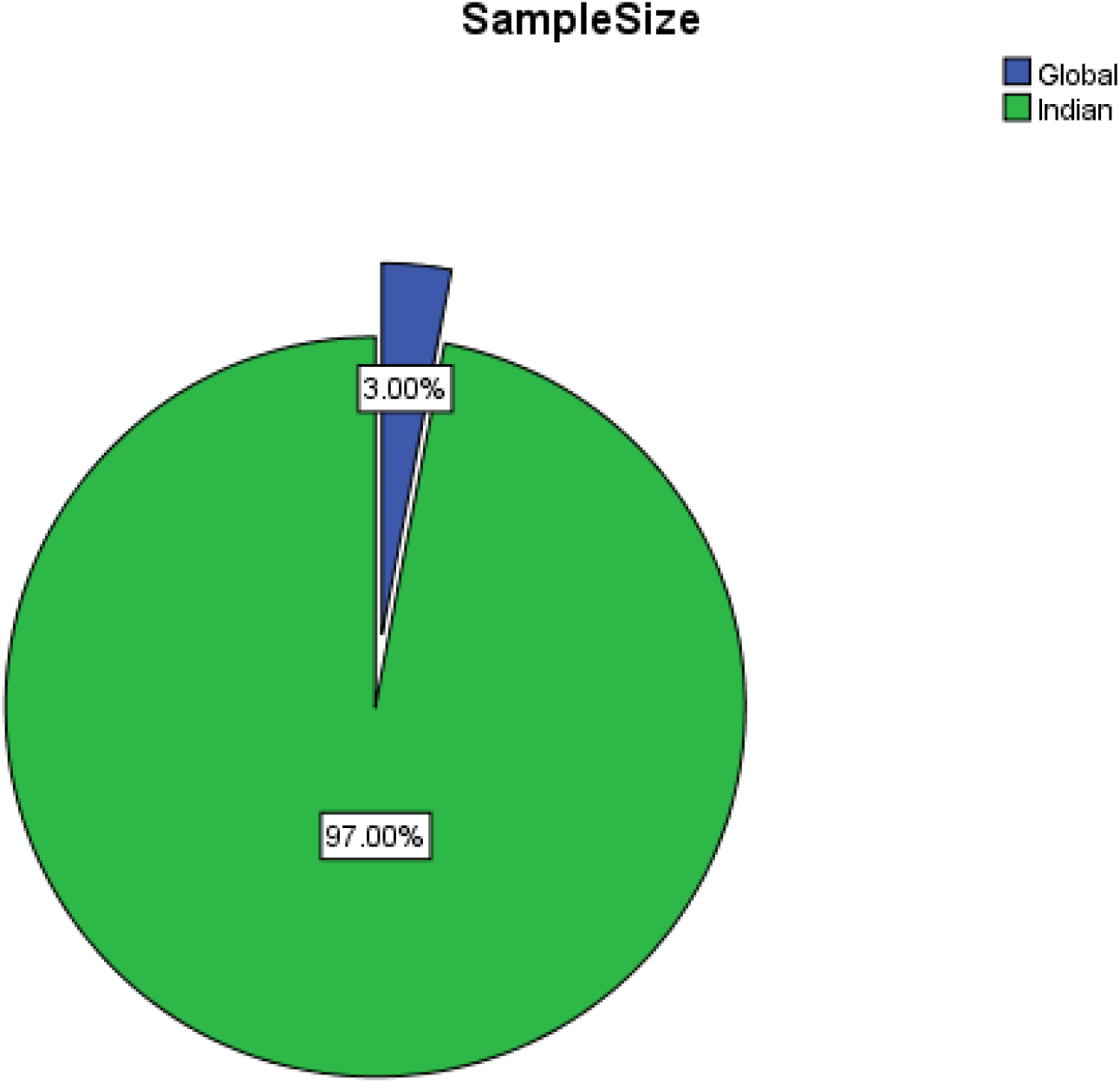
Sample Size.

### Sample size

The sample size varies between n=1 to n=5000 and above. 43% of trials have a sample size within 100 and followed by 32% of cases with a sample size within 500. This is followed by 11% of trials having a Sample size between 1 and 5000. 7% of trials have a large sample size above 5000, and 6% recruit within 1000 (Table – 1) (Figure -7).

**Figure: 7.**
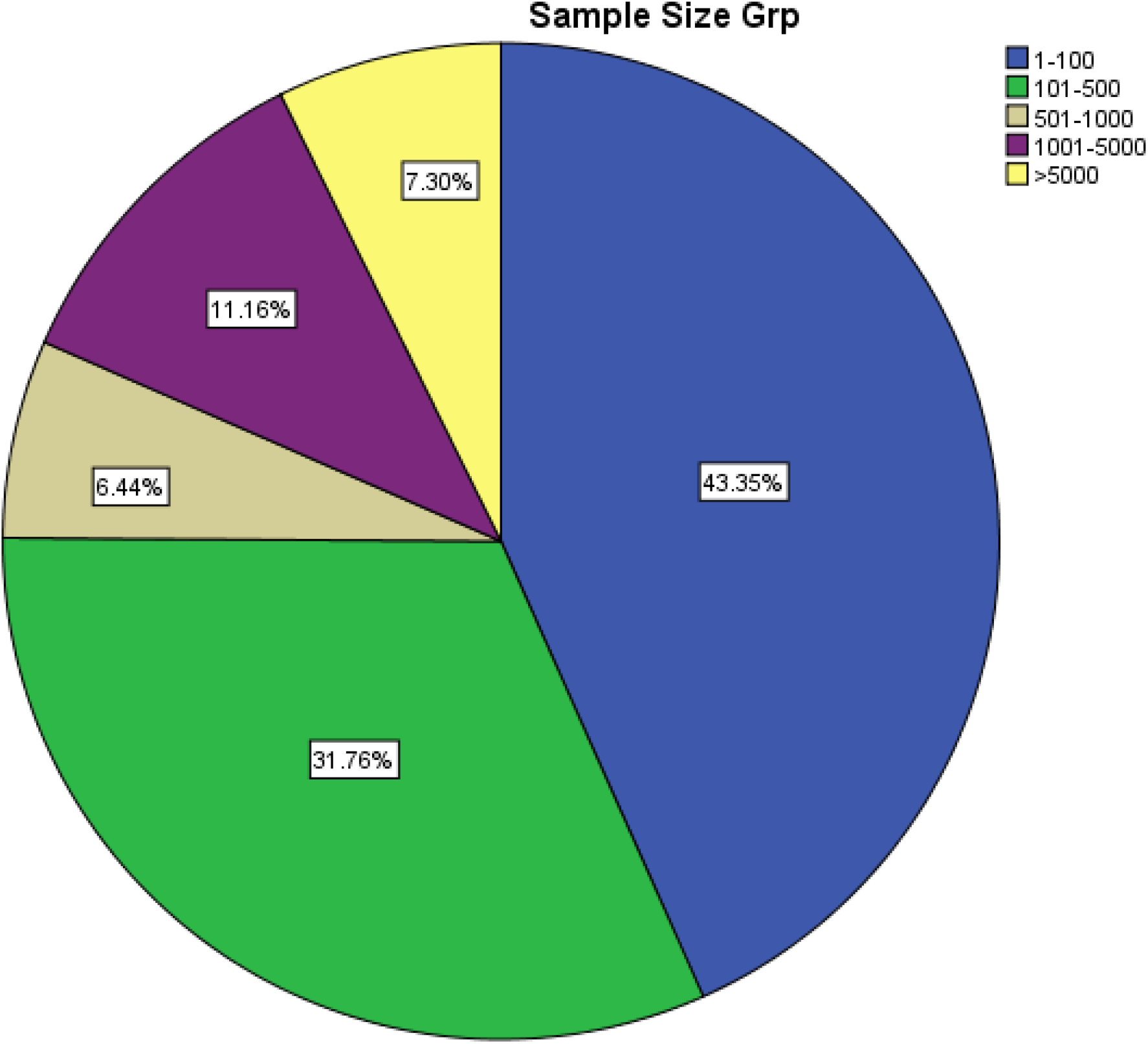
Sample Size Group.

### Randomization

Certain trials have adopted Adaptive Randomization, which is 3%, which has a chance of allocating patients following the already existing treatment groups. Stratified and Block Randomization occupies equal share (4% each). Stratified randomization helps in small trials, Permuted block randomization balances between the two intervention groups. However, many of the COVID 19 trials have been used—computer-generated randomization to prevent bias and human error. Computer Randomization occupies a significant share of 22% (Table – 1).

### Blinding

While 86% of Trials are not blinded, Double-blind trials are 5.5%, followed by Single blinded 5.15%. 1.72% of trials are triple blinded, and two trials are quadruple blinded (Table – 1).

### Trial Phases Involved

From the applicable 48%, below 4% evaluate the Safety of Drugs, Therapeutic exploratory is 24%, and Therapeutic confirmatory occupies 16%. Some trials have been listed as Phase IV supporting repurposing phenomena like Lozenges.

### Trial Duration

Five % of trials only showed trial duration greater than one year. 35% of both category with 1-3 month and 3 to 6 months falls in this duration (Figure -8).

**Figure: 8.**
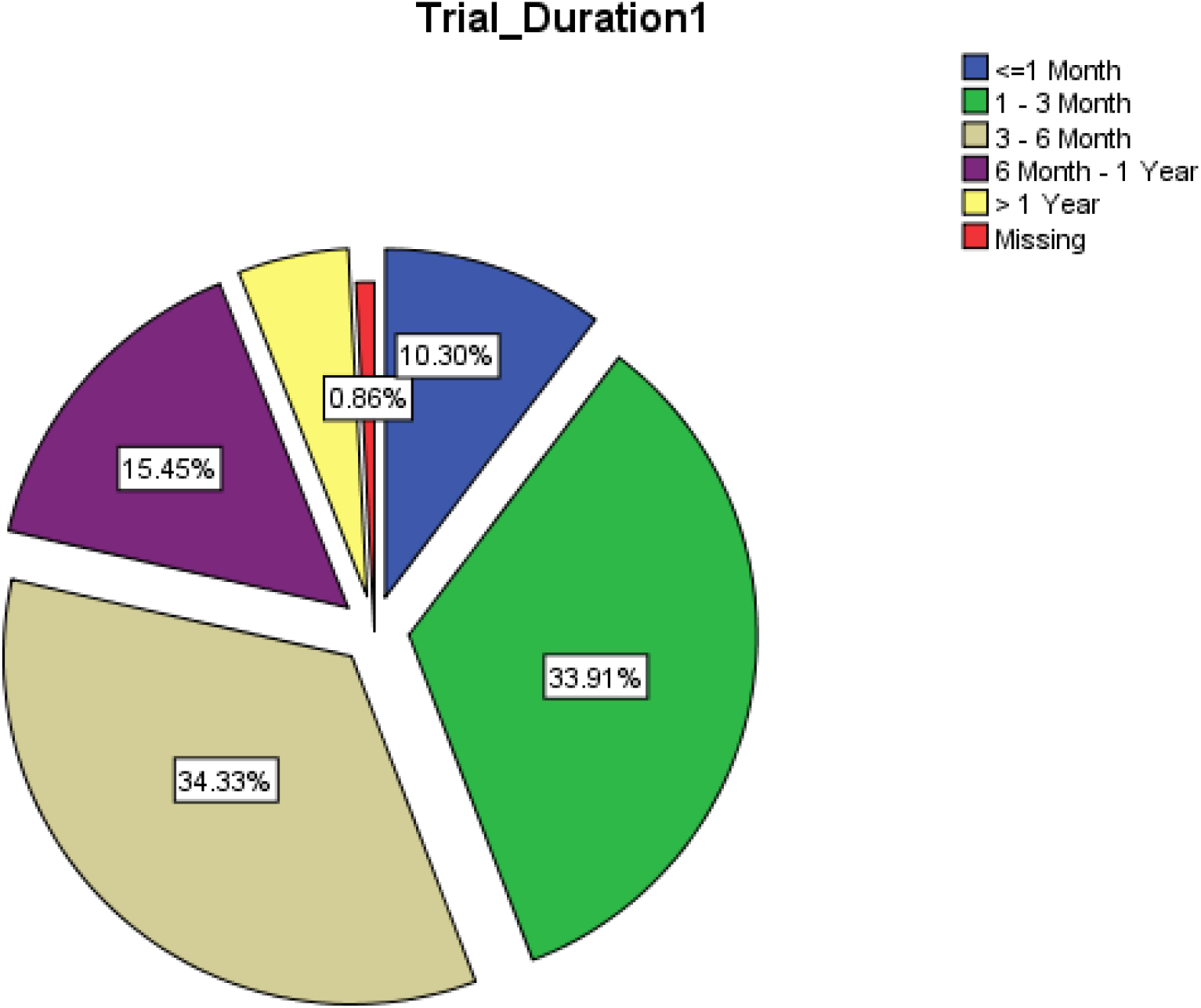
Trial Duration.

## Discussion

This study aims to measure the preparedness of various Indian health stakeholders controlled by two Ministries, one dealing with the conventional therapy (Ministry of Health and Family welfare) and the other with Indian traditional medicine (Ministry of AYUSH). Clinical trial registries are depots of newer ideas and newer interventions. The kinesis of conducting a Clinical trial during a pandemic is different from the normal situation. Understanding this scenario, Govt. of India has directed every clinical trial registered in CTRI. Ministry of AYUSH has released a particular GO, regulating the clinical trial during COVID-19. A bird-eye view and the Interventions used to support an ideology of repurposing existing drugs for novel coronavirus. PM of India has emphasized using immunomodulators from the AYUSH sector in his famous Mann Ke Baat talk. This study revealed that large-scale prophylactic prevention trials have been registered and used as a weapon to contain the disease to its might. Encouragingly, many medical colleges have collaborated with the AYUSH sector to integrate with the public’s interventions or compare the ordinary person’s benefits. It is evident that both healthy volunteers (32%) and affected individuals (68%) have also been subjected to trial in this pandemic situation, and many have consented to participate. It shows the tendency of humankind to find a solution to hunting problems and returning to normalcy.

Allopathic versus AYUSH interventions are equal in which Allopathy and Ayurveda occupy 49%, followed by Homeopathy, Siddha, Unani, and Yoga and Naturopathy in descending order.

As this a newer pandemic, after 100 years, a country like India and there exists a particular need for a keen understanding of the disease condition and its epidemiological aspects. This is reflected in the CTRI having a sizeable number of survey questionaries. Almost all the systems Ayurveda and Siddha, in particular, have integrated with Allopathy. Siddha, as integrated with Yoga and Ayurveda, has integrated with Homeopathy in certain hospitals.

At this pandemic, health stakeholders have taken a significant step to stick with standards reflected in 38% of the RCTs in CTRI. Next occupies open-ended single-arm trial followed by Non-randomized trial. It is tougher to follow a group in a pandemic, evident from only 0.5 % of cohort studies.

Research institutions and Medical colleges occupy a significant part of the trial, but only 10% of the pharmaceutical industry is taking up this issue. There exist much scope for pharmaceutical industries to invest more in real research. Global integrated trials are very minimal, which once again is an alarming sign in research.

Therapeutic exploratory and therapeutic confirmatory occupies a significant share while a sizeable number of trials have searched for Safety; based on the repurposing idea, some trials are designed as Phase IV, which are like measuring the benefit of lozenges among COVID-19 patients. The disease’s complexity has affected blinding in the trials, where only 14% of the trials are blinded. Among them, double-blinding occupies many, and some are triple and quadruple blinding.

At a given point in time, India has 107 trials than China, which expresses the country preparedness following the incidence and intensity of the disease.

## Conclusion

Despite the lockdown, the front line workers and health stakeholders of this country are serious about their business finding a solution for the problem stated, i.e., COVID-19. Allopathy and AYUSH’s health sectors have attempted in rapidity, which shows their surge to act against COVID-19. Revalidated interventions that have been used earlier in Dengue, Malaria, Immunomodulators and Earlier anti-virals and drugs used in Cancer and HIV are among the selected interventions. The outcome objectives were symptom reduction, getting negative in RT-PCR, reduction in hospitalization, minimal use of ICU, ventilators, and reduced mortality were the objectives observed. Post COVID-19 management clinical trials were not registered during the said time. However, there exists a lot of scope and necessity to do trials in Post Covid prospectively.

## Limitations

In this, the analysis was limited to a cross-sectional study of trials registered in the COVID-19 pandemic concerning AYUSH and Allopathy stream on CTRI. Like study type, study design, sponsorship, and relevant details on trial characteristics presented and analyzed from 1^st^ March 2020 to 22^nd^ June 2020. We did not include trials from the WHO database, US Canada, China, and other Clinical Trials Database. This analysis will give useful data based on CTRI by gathering information to improve quality research opportunities. In the future, follow-up is necessary to update results based on the information available to work further inconsistency.

## Data Availability

Will provide data on request

## Notes

### Competing Interest Statement

The authors have declared no competing interest.

### Author Declarations

Exempted from IRB

